# Prognostic accuracy of MALDI mass spectrometric analysis of plasma in COVID-19

**DOI:** 10.1101/2020.10.01.20205310

**Authors:** Lucas Cardoso Lazari, Fabio De Rose Ghilardi, Livia Rosa-Fernandes, Diego M Assis, José Carlos Nicolau, Veronica Feijoli Santiago, Talia Falcão Dalçóquio, Claudia B. Angeli, Adriadne Justi Bertolin, Claudio R. F. Marinho, Carsten Wrenger, Edison Luiz Durigon, Rinaldo Focaccia Siciliano, Giuseppe Palmisano

**Affiliations:** Department of Parasitology, Institute of Biomedical Sciences, University of São Paulo, São Paulo, Brazil; Instituto de Medicina Tropical, University of São Paulo, São Paulo, Brazil; Bruker do Brasil, Atibaia, São Paulo, Brazil; Heart Institute (InCor), University of São Paulo Medical School, São Paulo, Brazil; Department of Microbiology, Institute of Biomedical Sciences, University of São Paulo, São Paulo, Brazil

**Keywords:** COVID-19, SARS-CoV-2, Mass spectrometry, Biomarker, Plasma, Prognosis

## Abstract

**Purpose:** SARS-CoV-2 infection poses a global public health problem. There is a critical need for improvements in the noninvasive prognosis of COVID-19. We hypothesized that matrix-assisted laser desorption ionization mass spectrometry (MALDI-TOF MS) analysis combined with bottom-up proteomic analysis of plasma proteins might identify features to predict high and low risk cases of COVID-19.

**Patients and Methods:** We used MALDI-TOF MS to analyze plasma small proteins and peptides isolated using C18 micro-columns from a cohort containing a total of 117 cases of high (hospitalized) and low risk (outpatients) cases split into training (n = 88) and validation sets (n= 29). The plasma protein/peptide fingerprint obtained was used to train the algorithm before validation using a blinded test cohort.

**Results:** Several sample preparation, MS and data analysis parameters were optimized to achieve an overall accuracy of 85%, sensitivity of 90%, and specificity of 81% in the training set. In the blinded test set, this signature reached an overall accuracy of 93.1%, sensitivity of 87.5%, and specificity of 100%. From this signature, we identified two distinct regions in the MALDI-TOF profile belonging to the same proteoforms. A combination of 1D SDS-PAGE and quantitative bottom-up proteomic analysis allowed the identification of intact and truncated forms of serum amyloid A-1 and A-2 proteins. Conclusions: We found a plasma proteomic profile that discriminates against patients with high and low risk COVID-19. Proteomic analysis of C18-fractionated plasma may have a role in the noninvasive prognosis of COVID-19. Further validation will consolidate its clinical utility.

**Key message:** *What is the key question?:* Do individuals infected with SARS-CoV-2 harboring different degree of disease severity have a plasma protein profile that differentiate them and predict the COVID-19 outcome?

*What is the bottom line?:* In a series of 117 patients with COVID-19 divided in hospitalized (60) and outpatients (57), differential expression of serum amyloid A-1 (SAA1) and A-2 (SAA2) predict their outcome.

*Why read on?:* The high mortality rate in SARS-CoV-2 infected individuals requires accurate markers for predicting COVID-19 severity. Plasma levels of SAA1 and SAA2 indicate higher risk of hospitalization and can be used to improve COVID-19 monitoring and therapy.

## Introduction

The pandemic of SARS-CoV-2 infection, the etiological agent of coronavirus disease 2019 (COVID-19), has affected millions of people worldwide. The first case was reported in Wuhan, China and as for September 30^th^, 33,722,075 people have been infected and 1,009,270 died. The ongoing outbreak is considered a pandemic (World Health Organization). The symptoms range from mild with fever, dry cough, headache, fatigue and loss of taste and smell to severe complications including difficulty breathing or shortness of breath, chest pain and loss of speech or movement that can lead to hospitalization and death.^1^ Although vaccines and small molecule treatments are in clinical trial, no definitive treatment for COVID-19 is available yet.^2–4^ A mortality rate of approximately 4% has been detected in COVID-19 patients compared to 0.1% in influenza infection (World Health Organization). Due to that, it is imperative to identify patients at high risk for severe illness to assist them with supportive therapy. Markers of COVID-19 severity have been proposed.^5–7^

MALDI-MS has been successfully implemented into the microbiology field building reference spectral libraries for rapid, sensitive and specific identification of bacterial and fungal species.^8^ This approach is well established and accepted in many countries for routine diagnostics of yeast and bacterial infections. Viral species identification has been elucidated using similar strategies.^9^ Recently, MALDI-TOF MS analysis of nasal swabs allowed sensitive and specific detection of SARS-CoV-2 infection.^10^ Moreover, MALDI-TOF MS analysis of human biofluids have been proposed as diagnostic and prognostic techniques in several diseases ranging from cancer, cardiovascular, neurological and infectious diseases.^11–15^

This study shows the identification of a plasma proteomic signature obtained from high (hospitalized) versus low (outpatients) risk patients with COVID-19 using an easy to perform, rapid and widespread technology such as MALDI-TOF MS, present in several clinical laboratories worldwide. A training and validation dataset allowed the prioritization of discriminant features identified using bottom-up quantitative proteomics. SAA-1 and SAA-2 proteoforms were differentially expressed between the two groups allowing the implementation of point-of-care diagnostics. More studies, including larger inter-institutional cohorts, are needed to move this marker into the clinic.

## Materials and Methods

### Study Subjects and Design

Plasma from a total of 117 patients with COVID-19 divided into high risk (n = 57) and low risk (n = 60) was collected prospectively from a Brazilian cohort (**Tables 1 and 2**) at the Heart Institute (InCor) and Central Institute, University of São Paulo Medical School, Brazil between March 2020 to July 2020 (CAAE 30299620.7.0000.0068). Patients with high risk were defined based on clinical parameters evaluated at the time of admission that required hospitalization compared to low risk patients. Cases were included with a clinical picture suggestive of COVID-19 defined as two or more of the following: cough, fever, shortness of breath, diarrhea, myalgia, headache, sore throat, running nose, sudden gustatory or olfactory loss and detection of viral RNA in nasopharyngeal SARS-CoV-2 PCR positive. Patients with high and low risk of hospitalization were matched for confounding variables such as age, sex, and co-morbidities to explain the difference between groups (**Supplementary Table 1**). The matched groups were split into a training (n=88) and a validation set (n=29). A proteomic signature obtained by MALDI-TOF MS analysis in a training set was tested in the validation set of matched groups. A total of 75% of the plasma samples were assigned to a training set and the remaining to a validation test set. Training and validation sets were matched according to the same criteria (**Table 3**). Plasma were collected, aliquoted and stored at −80°C for further analyses.

**Table 1.**
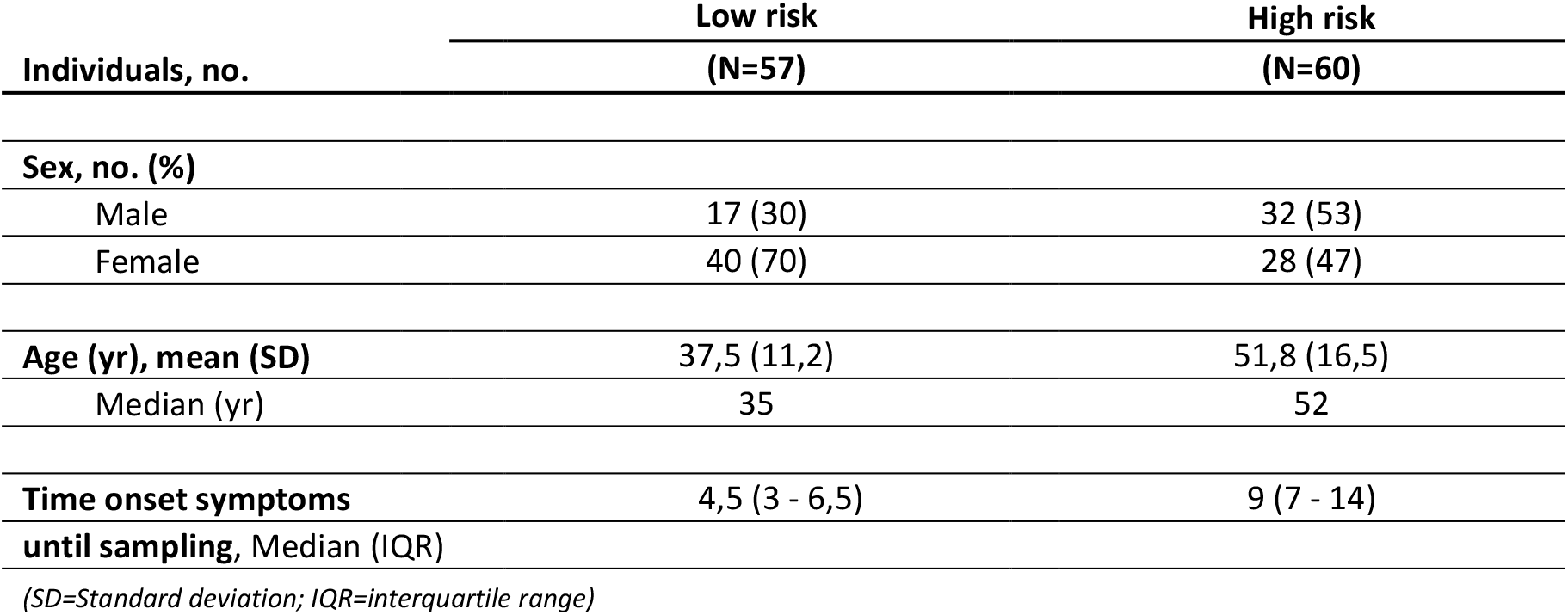
Epidemiological characteristics of the 117 COVID-19 patients investigated in this study.

**Table 2.**
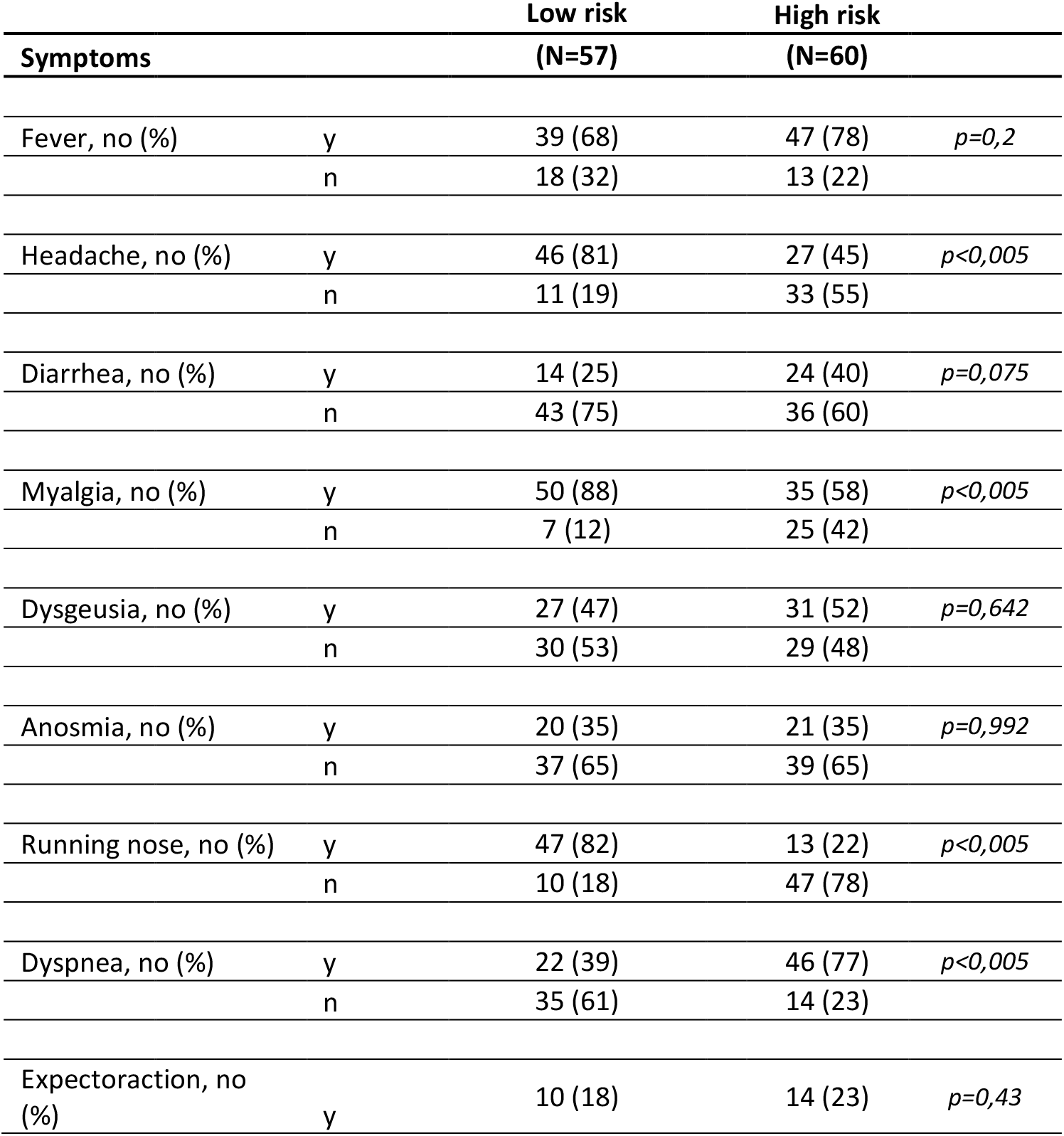
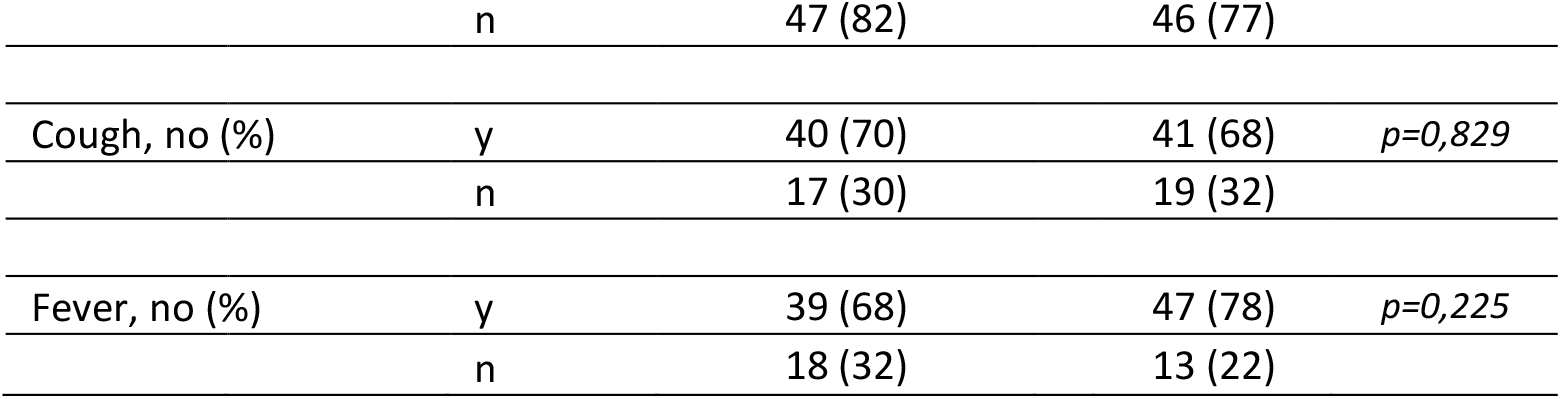
Clinical findings associated with the 117 COVID-19 patients investigated in this study.

| Symptoms |  | Low risk<br>(N=57) | High risk<br>(N=60) |  |
| --- | --- | --- | --- | --- |
| Fever, no (%) | y | 39 (68) | 47 (78) | $p=0,2$ |
|  | n | 18 (32) | 13 (22) |  |
| Headache, no (%) | y | 46 (81) | 27 (45) | $p<0,005$ |
|  | n | 11 (19) | 33 (55) |  |
| Diarrhea, no (%) | y | 14 (25) | 24 (40) | $p=0,075$ |
|  | n | 43 (75) | 36 (60) |  |
| Myalgia, no (%) | y | 50 (88) | 35 (58) | $p<0,005$ |
|  | n | 7 (12) | 25 (42) |  |
| Dysgeusia, no (%) | y | 27 (47) | 31 (52) | $p=0,642$ |
|  | n | 30 (53) | 29 (48) |  |
| Anosmia, no (%) | y | 20 (35) | 21 (35) | $p=0,992$ |
|  | n | 37 (65) | 39 (65) |  |
| Running nose, no (%) | y | 47 (82) | 13 (22) | $p<0,005$ |
|  | n | 10 (18) | 47 (78) |  |
| Dyspnea, no (%) | y | 22 (39) | 46 (77) | $p<0,005$ |
|  | n | 35 (61) | 14 (23) |  |
| Expectoration, no (%) | y | 10 (18) | 14 (23) | $p=0,43$ |
|  | n | 47 (82) | 46 (77) |  |
| Cough, no (%) | y | 40 (70) | 41 (68) | $p=0,829$ |
|  | n | 17 (30) | 19 (32) |  |
| Fever, no (%) | y | 39 (68) | 47 (78) | $p=0,225$ |
|  | n | 18 (32) | 13 (22) |  |

**Table 3.**
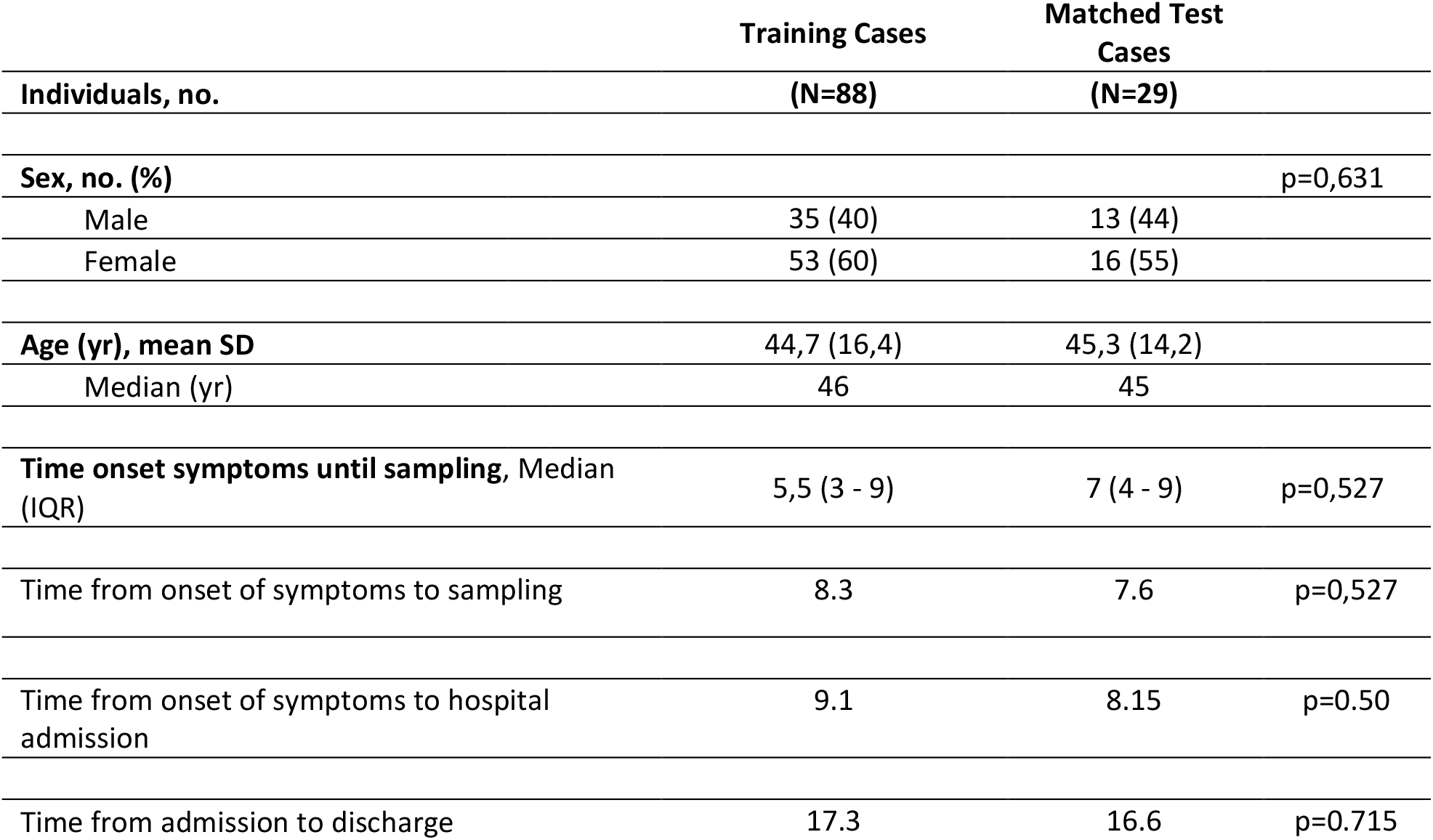
Epidemiological Characteristics in Training and Test COVID 19 datasets

|  | Training Cases | Matched Test Cases |  |
| --- | --- | --- | --- |
| Individuals, no. | (N=88) | (N=29) |  |
| **Sex, no. (%)** | | | $p=0,631$ |
| Male | 35 (40) | 13 (44) |  |
| Female | 53 (60) | 16 (55) |  |
| **Age (yr), mean SD** | 44,7 (16,4) | 45,3 (14,2) |  |
| Median (yr) | 46 | 45 |  |
| **Time onset symptoms until sampling, Median (IQR)** | 5,5 (3 - 9) | 7 (4 - 9) | $p=0,527$ |
| Time from onset of symptoms to sampling | 8.3 | 7.6 | $p=0,527$ |
| Time from onset of symptoms to hospital admission | 9.1 | 8.15 | $p=0.50$ |
| Time from admission to discharge | 17.3 | 16.6 | $p=0.715$ |

Sample preparation for MALDI-TOF MS, LC-MS/MS analysis and data processing details are provided in the **Supplementary methods**.

## Results

### Method optimization and evaluation of reproducibility and variability

The analytical platform shown in this study was developed through three phases: 1) MALDI-TOF MS-based method development for plasma samples, 2) clinical application to plasma isolated from COVID-19 patients with high and low risk and 3) identification of markers to discriminate high and low risk patients, according to the experimental workflow (**Figure 1**).

**Figure 1:**
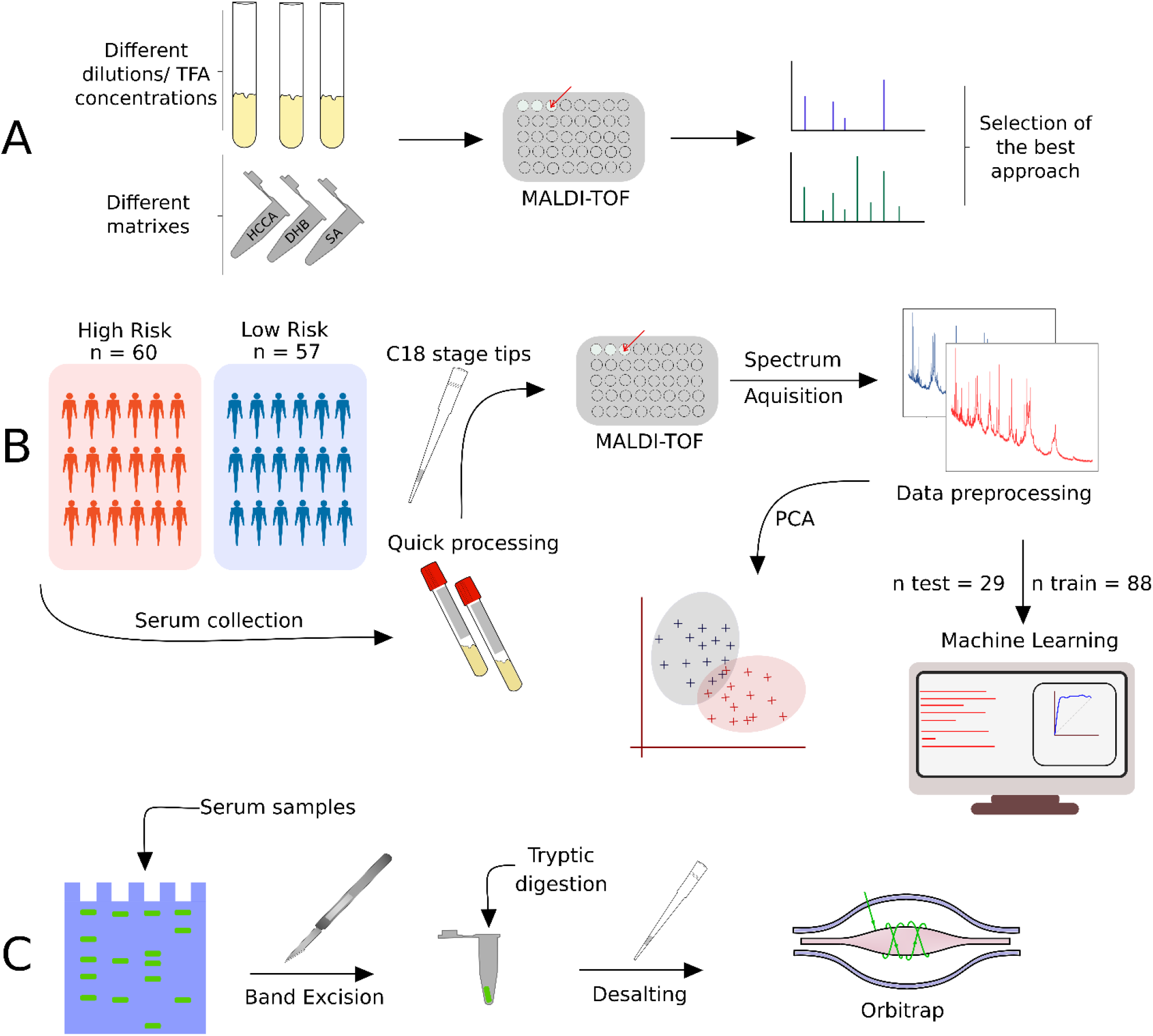
Experimental workflow applied to this study. A) Method development for MALDI-TOF MS analysis of plasma samples. B) MALDI-TOF MS analysis of 117 COVID-19 patients combined with machine learning to identify MS discriminant features in the training and test dataset. C) Biomarker discovery based on 1D SDS-PAGE and nLC-MS/MS analysis.

Initial method development focused on the selection of the appropriate matrix using unfractionated plasma. The dried droplet sample preparation method using unfractionated plasma and three matrices (HCCA, DHB and SA) was tested acquiring the protein/peptide profile in automatic mode. Using the HCCA matrix resulted in the detection of more peaks compared to other matrices (**Supplementary Figure 1**). The highest peaks at 16616.3, 13315.1, 11095.6, 9496.2 and 8316.8 m/z corresponded to human serum albumin with 4-8 charges. To improve the number of peaks detected, the acid concentration within the matrix (TFA 2.5%) was increased (**Supplementary Figure 2**). The peak intensity increased and two peaks in the 6000 m/z region were detected; however, the serum albumin peaks were still within the most abundant. In order to improve the number of peaks detected for each spectrum, C18-based plasma fractionation was performed. The MALDI-TOF performances were evaluated measuring the number of peaks and the variation in intensity and frequency of specific peaks detected for each spectrum after processing as described in the Materials and Methods. A total of 5 plasma were tested. C18-based fractionation showed a higher number of peaks detected from 2000-10000 m/z than unfractionated plasma (**Supplementary Figure 3**). A comparison between three matrices showed that HCCA yielded a higher number of peaks and intensity than other matrices (**Supplementary Figure 4 and 5**).

Due to that, we chose to perform all analyses using a C18 fractionated sample eluting the proteins/peptides from the microcolumn using the HCCA matrix containing 50% acetonitrile and 2.5% TFA as described in the materials and methods section. To test if sample pretreatment induces potential artifacts that affect the reproducibility of the entire strategy, quadruplicate analysis of the same spot and analysis of each sample on three different preparations was performed (**Supplementary Figure 6**). Two m/z regions were selected to calculate the coefficient of variance (CV) considering the sample preparation and MS acquisition variability. Within the 5700-5900 m/z region, an average of 10% CV was obtained. Within the 11300-11700 m/z region, an average of 20% CV was obtained (**Supplementary Figure 7**). The lower CV in the higher m/z region is associated with the lower intensity, which increases the variance within and between samples. The CVs data obtained are in agreement with other reports using MALDI-TOF MS profiling of biofluids.^16–18^ Due to that, the optimized sample preparation strategy applied to this study was based on 1ul of plasma fractionated using C18 microcolumns and proteins/peptides were eluted using HCCA matrix containing 2.5% TFA.

### Prognostic value of the Plasma proteome profile of COVID-19 patients

In the next phase, we applied the optimized strategy to plasma samples collected from COVID-19 patients. 117 patients with laboratory-confirmed COVID-19 disease were enrolled in this study, 57 with mild disease, no need for hospitalization (low risk) and 60 being admitted in the hospital (high risk). Their status was assessed using a combination of molecular, serological and clinical examination. RT-PCR and ELISA were used to test the active or past SARS-CoV-2 infection. **Table 1** shows the demographic characteristics of these two groups of patients. The median age was significantly higher in the hospitalized group (52 years; IQR 39,5 - 64,5) than the mild group (35 years; IQR 29 - 47). A total of 40 (70%) of outpatients were female and only 28 (47%) were female in the hospitalized group. The median time of symptom onset before blood sampling was also higher in the severe group (9 days and IQR 7 - 14) than in the outpatients group (4,5 days and IQR 3 - 6,5 using two-tailed Mann–Whitney U-test). Four patients (3,5%) died (**Supplementary Figure 8**). The sex and age distribution observed in this study is in line with the literature findings. Indeed, a significant association between sex, age and COVID-19 disease prognosis has been reported.^19^ Male patients have a higher mortality rate, hospitalizations and lower chance of recovery compared to females.^19–21^ It has been shown that female patients have higher plasma levels of IL-8 and IL-18 cytokines and different immune cells number and type sustained along the life that reduce the severity of COVID-19.^22^ The most prevalent symptoms in this cohort were fever and myalgia, in patients that were hospitalized with dyspnea (77%) and cough (68%) were the main clinical features. In the group of outpatients, upper respiratory signs as rhinorrhea (82%) and headache (81%) were more prevalent (**Table 2**). The difference between groups was statistically significant (chi-square test p<0,005) regarding the type of symptom presented: as rhinorrhea (82% of the mild symptomatic patients and 22% of admitted in hospital patients), headache (81% of the mild symptomatic and 45% of hospitalized patients) and myalgia (88% of the mild symptomatic and 58% of hospitalized patients) being more prevalent in patients not requiring hospitalization. On the other hand, dyspnea was present in only 22% of the mild symptomatic patient and in 77% of the patients that were admitted in a hospital (p<0,005). Patients with a mild presentation of COVID-19 (low risk) presented running nose, headache and myalgia and did not develop the inflammatory syndrome. This group of patients were more attentive to their first symptoms and search for health care earlier (most of them are healthcare professionals which may represent a bias). The most prevalent comorbidities found in our patients were obesity (4% of the mild group and 27% of the hospitalized group) and dyslipidemia (5% of the mild group and 17% of the hospitalized group). Although there was no difference in dyslipidemia’s prevalence between both groups we found a higher number of obese patients (BMI > 30) between people requiring hospitalization (16/60 = 27%), **Supplementary Table 1**. It is important to mention that this survey was done at INCOR hospital, an institution specialized in heart and lung diseases, so we did find a higher prevalence of cardiovascular diseases than in the general population (as heart transplantation patients and people with chronic conditions). MALDI-TOF spectra obtained from the C18 fractionated plasma from the high and low risk groups were analyzed, as described in the Materials and Methods section. Analyzing the 117 plasma samples, the data preprocessing yielded a total of 63 peaks detected, which dropped to 38 after the Wilcoxon rank sum test with a p-value < 0.05. After Ig filtering, 5 total peaks were identified (**Supplementary Table 2**). PCA analysis of significant peaks and Ig filtered peaks are presented in **Figure 2**. The Ig filtering demonstrated a better separation than the PCA with all significant peaks. The MS peaks were analyzed using six machine learning algorithms to discriminate between the two conditions under optimized parameters (**Supplementary Table 3**). In general, all models did not differ significantly from each other, and presented a robust behavior comparing each fold. The results of the six tested models in the dataset without Ig filtering showed that the RF model had the higher mean area under the curve (AUC) for the ROC curve (0.95) (**Figure 3A**); additionally, SVM-P and RF had the lowest standard deviation for AUC values between each fold (**Supplementary Table 4, 5**). Thus, the RF model was considered to have the best performance of the six tested and was applied to the test set. For Ig filtering approach, RF had the highest mean AUC for ROC (0.94) with a low standard deviation between each fold (**Figure 3B**); so it was considered the model with the best performance for Ig filtered peaks. The best parameter for RF is mtry=2 for both. It is worth mentioning that KNN, NNET, SVM-P and SVM-R also had good performances. The ROC and PR curves of the training set for Ig filtered and non-filtered peaks, together with the mean accuracy, mean sensitivity and mean specificity of the prediction in the validation set in each fold (**Figure 3C and D**).

**Table 4.** Sequences of truncated serum amyloid protein A-1 and A-2 identified as discriminant peaks in the MALDI-TOF MS analysis and sequenced using nLC-MS/MS.

| Protein name | Sequence | MW,<br>experimental<br>(Da) | MW,<br>theoretical<br>(Da) |
| --- | --- | --- | --- |
| Serum amyloid A-1<br>(SAA1) | <sup>19</sup>RSFFSFLGEAFDGARDMWRAYS---AGLPEKY<sup>122</sup> | 11683 | 11675.49 |
| Serum amyloid A-1<br>(SAA1) | <sup>20</sup>SFFSFLGEAFDGARDMWRAYS---AGLPEKY<sup>122</sup> | 11530 | 11519.39 |
| Serum amyloid A-1<br>(SAA1) | <sup>21</sup>FFSFLGEAFDGARDMWRAYS---AGLPEKY<sup>122</sup> | 11443 | 11432 |
| Serum amyloid A-2<br>(SAA2) | <sup>19</sup>RSFFSFLGEAFDGARDMWRAYS---AGLPEKY<sup>122</sup> | 11633 | 11640.60 |
| Serum amyloid A-2<br>(SAA2) | <sup>20</sup>SFFSFLGEAFDGARDMWRAYS---AGLPEKY<sup>122</sup> | 11476 | 11484.50 |
| Serum amyloid A-2<br>(SAA2) | <sup>21</sup>FFSFLGEAFDGARDMWRAYS---AGLPEKY<sup>122</sup> | 11393 | 11397.47 |

**Figure 2:**
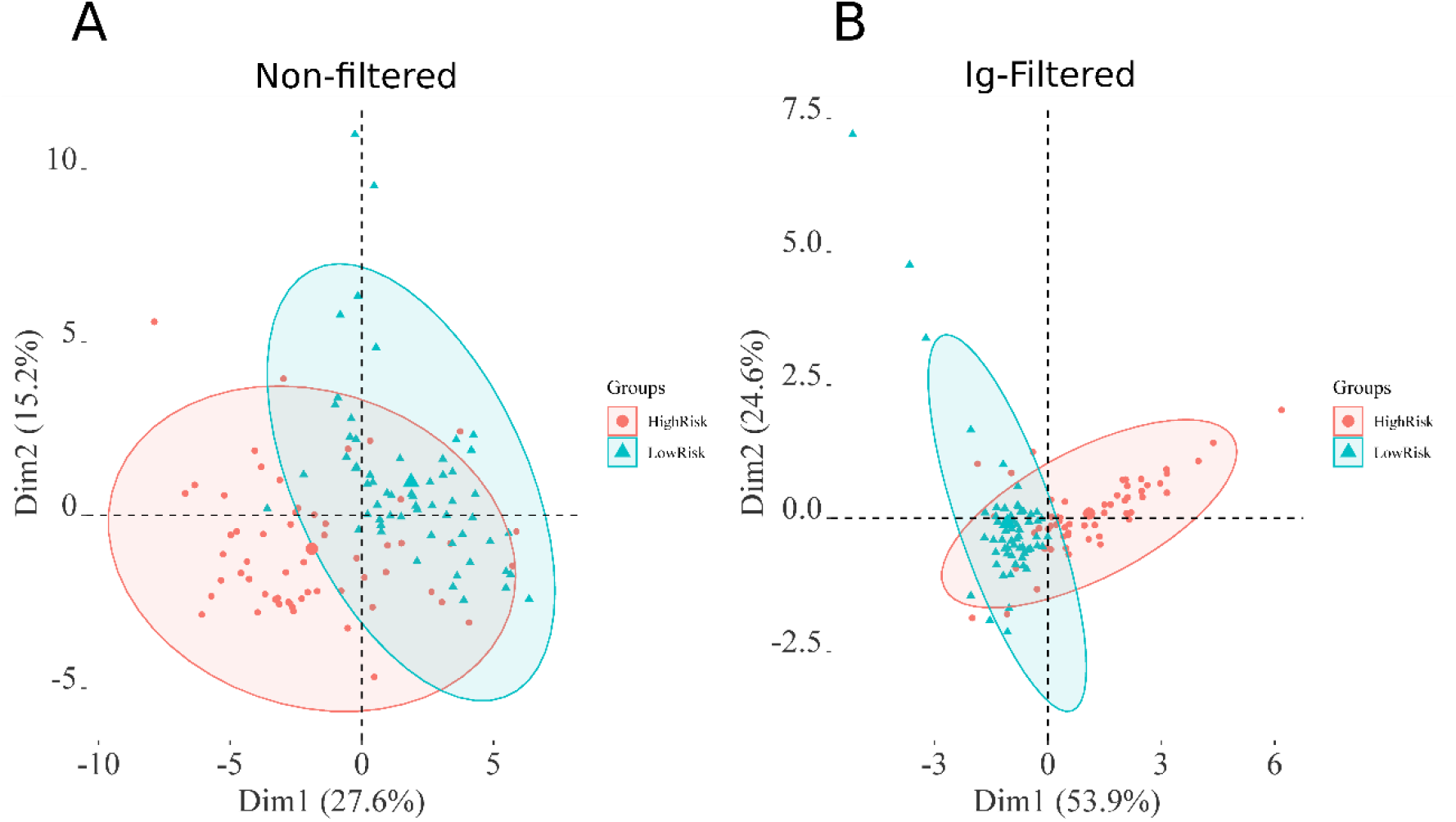
PCA analysis of the preprocessed MALDI-TOF MS spectra obtained from 117 plasma samples. A) PCA of all significant peaks. B) the PCA of peaks selected with the Ig (Information gain) method.

**Figure 3:**
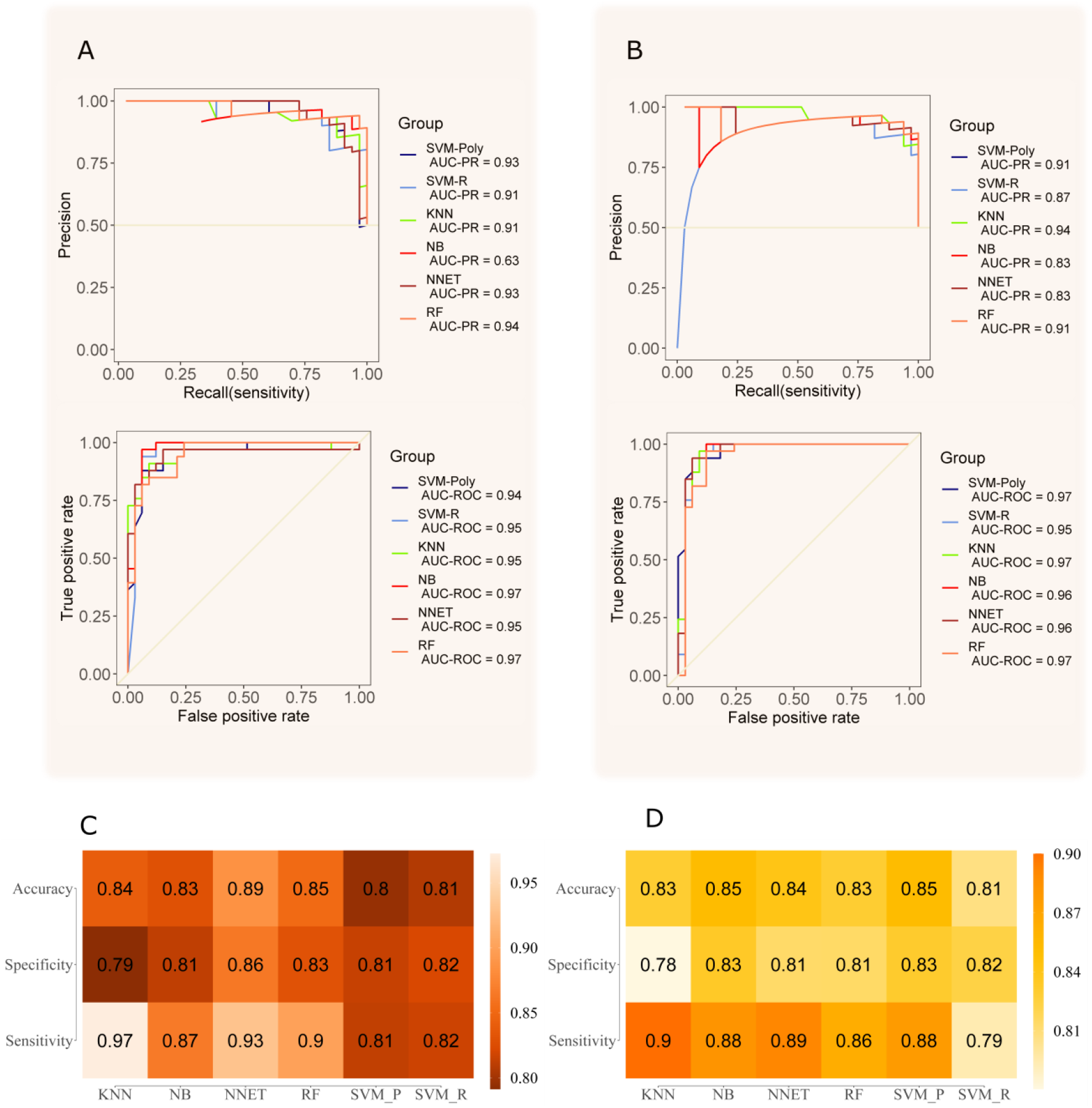
Four-fold nested cross validation of the training set for model selection. A) best AUC values for ROC and PR curves of each model with the non-filtered dataset. In B, best AUC values for ROC and PR curves of each model with the Ig filtered dataset. In C, the average accuracy, specificity and sensibility of all folds for non-filtered peaks. In D, the average accuracy, specificity and sensibility of all folds for Ig filtered peaks.

The best model with optimized parameters was applied to the test set. The ROC curves on this dataset without and with Ig selection resulted in 0.95 and 0.94 AUC, respectively. The PR curves applied on the same dataset without and with Ig selection resulted in 0.92 and 0.91 AUC, respectively (**Figure 4A and B**). Applying the RF model with optimized parameters to the confusion matrix gave an accuracy of 0.931 and 0.862 without and with Ig filtering (**Figure 4C, D and E**). The filtering by information gain demonstrated a lower performance when compared to the modeling without Ig filtering. However, in the model selection step, the data without this filtering showed an overfitting in one of the four folds, while the data that was filtered did not (**fold 3 of Supplementary Table 4**).

**Figure 4:**
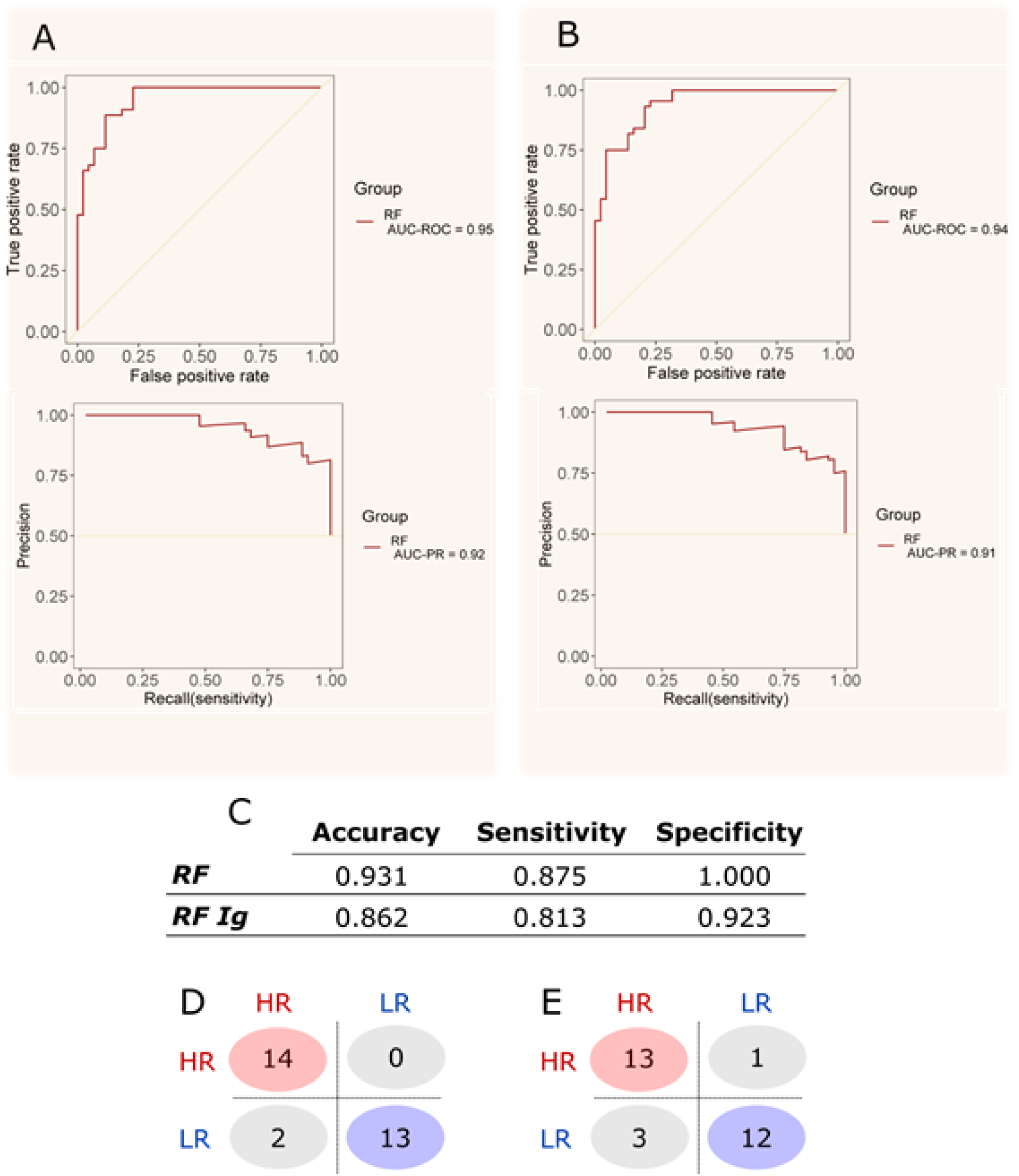
Training results of the model with best performance (RF) and the prediction results of the test set. In A and B boxes, the ROC and PR curves of the training step for the dataset without Ig selection and with Ig selection, respectively. C) the metrics of the confusion matrix from the test set prediction of both approaches.D) the confusion matrix of the prediction for the model without Ig filtering. E) the confusion matrix of the prediction for the model with Ig filtering.

### Biomarker identification

Next, we focused on the identification of specific biomarkers based on the MALDI-TOF profile obtained. Within the MS features with the highest discriminatory value, statistical rank, relative m/z peak intensity, a cluster of signals in the 5700-5900 and 11300-11700 m/z regions was chosen as a specific signature able to distinguish high from low risk COVID-19 patients (**Figure 5**). Interestingly, the peaks at 5696, 5724, 5739, 5765, 5818 and 5843 Da correspond to the doubly charged ions of 11393, 11443, 11476, 11530, 11633 and 11683 Da, respectively. This indicates that one or more proteoforms are contributing to discriminating high and low risk patients. To identify these proteins, the C18 fractionated plasma proteins/peptides were separated using 1D SDS-PAGE (**Figure 6**). The 10000-15000 Da region of the gel was excised, in-gel digested, and analyzed using nanoflow LC-MS/MS followed by data analysis. Quantitative proteomic analysis allowed the identification of 179 proteins with at least one unique peptide (**Supplementary Table 6 and 7**). Serum albumin, serotransferrin, complement C3a, alpha-2 macroglobulin and haptoglobin were among the proteins with the highest PSMs. These proteins have molecular weights (MW) higher than 15kDa, which is the MW cutoff of the gel band analyzed. A total of 52 proteins were identified with a MW between 10-15kDa. Within them, four proteins, platelet factor 4 (PF4), immunoglobulin lambda variable 4-69 (IGLV4-69), serum amyloid A-1 (SAA1) and serum amyloid A-2 (SAA2) were upregulated in the high compared to low risk groups. The MALDI-TOF MS peaks at 11443, 11530 11683 Da correspond to truncated fragments originating from serum amyloid A1 (**Table 3**). The MALDI-TOF peaks at 11633, 11476 and 11393 Da correspond to truncated fragments originating from serum amyloid A2 (**Table 3**). The peptide 20-SFFSFLGEAFDGAR-33 peptide, shared between SAA1 and SAA2, was identified in the LC-MS/MS analysis and constitutes the initial sequence of one truncated form. The R19 was cleaved by trypsin during processing and constitutes the first amino acid of the other truncated form. The 21-FFSFLGEAFDGAR-33 semi-tryptic peptide, shared between SAA1 and SAA2, was also identified and constitutes the initial sequence of the other truncated form (**Supplementary Table 7**). Moreover, the semi-tryptic peptides 23-SFLGEAFDGAR-33 and 24-FLGEAFDGAR-33 were identified, suggesting the presence of another two low abundant proteoforms, which were not detected in the MALDI-TOF MS spectra. These sequences were consistent with the molecular weights of the discriminatory m/z values identified upregulated in the plasma samples collected from COVID-19 patients with high risk.

**Figure 5:**
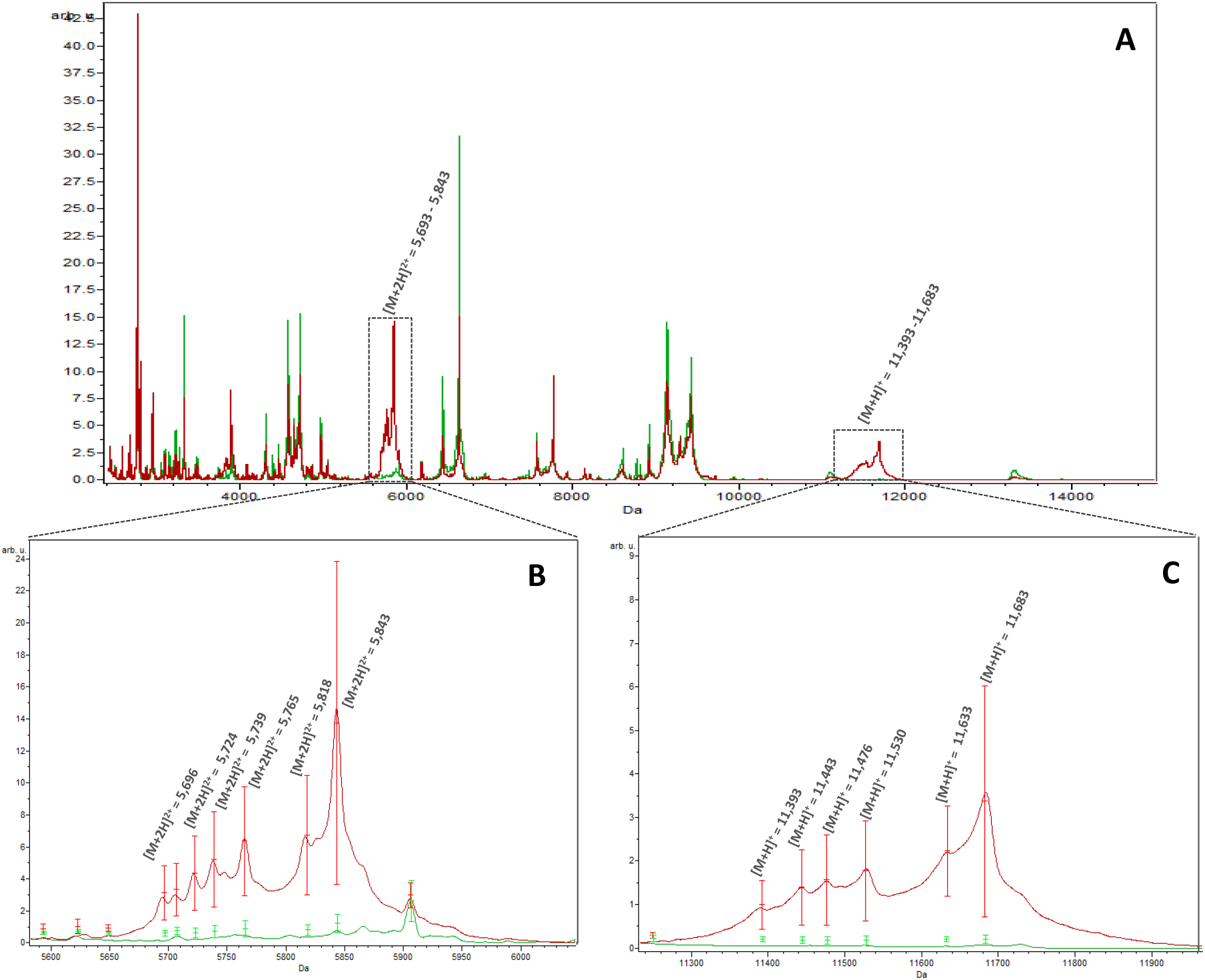
Average MALDI-TOF MS spectra obtained using C18 fractionated plasma collected from high risk (red) and low risk patients (green). (A) Full mass range, (B) zoom at 5845 m/z and (C) zoom at 11683 m/z. Standard deviations of intensities are represented by vertical lines.

**Figure 6:**
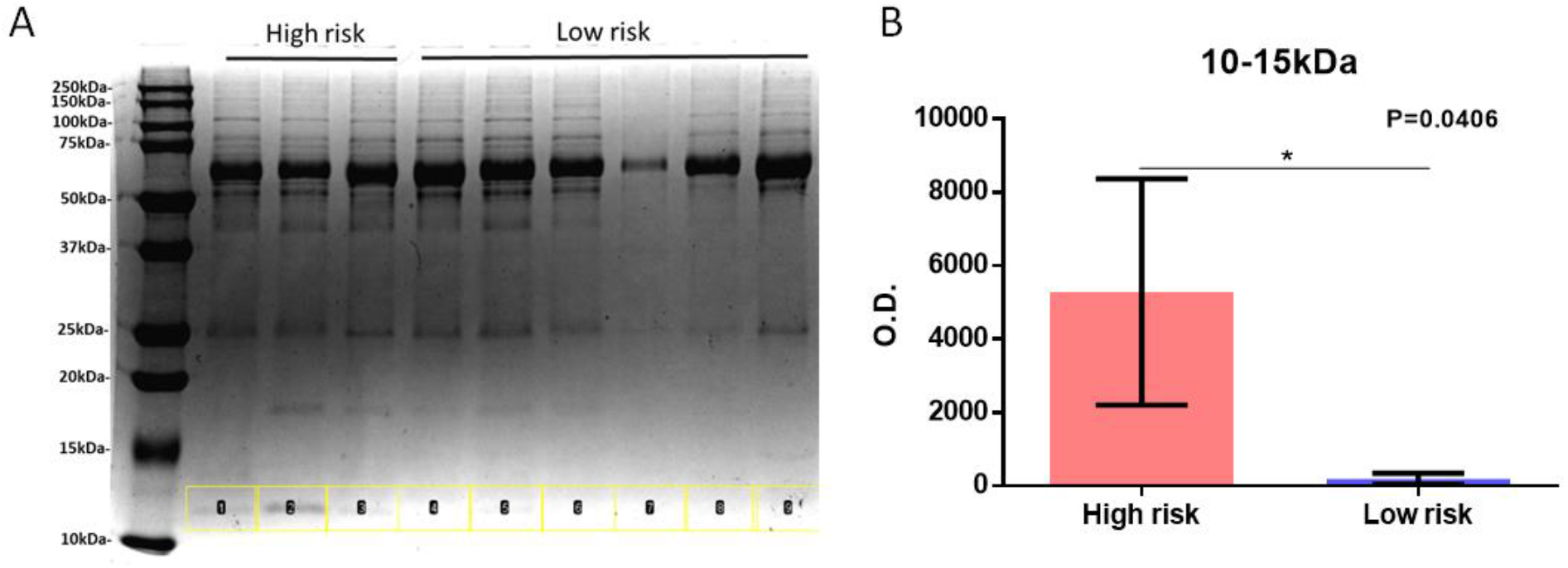
1D-SDS PAGE of C18 fractionated plasma from high (3) and low (6) risk patients. A) The region between 10-15kDa is highlighted in yellow. B) Quantification of the 10-15kDa region in the high and low risk groups.

## Discussion

We describe the application of MALDI-TOF MS to identify a protein signature specific to COVID-19 patients with high and low risk, based on clinical symptoms, using 1ul of C18 fractionated plasma. This study was based on the supposition that SARS-CoV-2 infection induces a systemic response that changes selectively the plasma protein expression, allowing a discrimination between patients at high risk (need of hospitalization) compared to low risk ones (outpatient treatment). Using machine learning algorithms with optimized parameters, an overall accuracy of 93.1%, a sensitivity of 87.5%, and a specificity of 100% was achieved separating the two groups. The sample preparation, data acquisition and analysis parameters were optimized and validated to understand their influence of these factors in creating systemic biases. We confirmed that these factors were not influencing the accuracy of our approach based on the CVs detected. CVs reported in this study confirm similar reports from other research groups.^17,23,24^ The overfitting issue, normally seen in these approaches, was minimized using independent datasets for the training and the validation. Moreover, the test dataset was analyzed only once based on the results from the training set avoiding subjective selection of the best results. In this study, we were interested in determining a specific protein or a panel of proteins that could be used for COVID-19 prognosis. Applying a combination of gel electrophoresis and nLC-MS/MS, we identified SAA1 and SAA2 proteoforms as regulated discriminatory proteins. These two proteins are involved in the acute phase response. Proteins involved in the acute phase response are increased early during viral and bacterial infections. Serum amyloid A-1 (SAA1) and A-2 (SAA2) are acute phase reactants synthesized by the liver and secreted into the bloodstream inflammatory and oncogenic processes.^25^ Extra-hepatic SAA protein synthesis has been reported in inflamed tissues.^26,27^ SAA represents a family of high-density lipoproteins with 103-104 amino acids sharing high sequence homology between the different members. Four isoforms are expressed in humans SAA1, SAA2, SAA3 and SAA4.^25,28^ During infection, SAA protein production and secretion in the circulation can increase more than 1000-fold suggesting an early response to infection. However, sustained expression of SAA proteins is associated with chronic pathological conditions. SAA1 was already reported to be differentially expressed in patients with severe.^29,30^ The SAA1 and SAA2 proteins were also identified upregulated in severe COVID-19 patients in a clinical cohort from China.^31^ The authors used large scale LC-MS/MS analysis of serum samples to identified differentially regulated proteins and metabolites as potential prognostic markers.^31^ The identification of SAA1 and SAA2 as potential markers confirms our study.A specific correlation between SAA proteins and CRP has been found in several infectious diseases with the concentration of SAA increasing up to 2000mg/L.^32^ However, SAA proteins were found to be more sensitive than CRP in detecting variation in the inflammatory status of infected patients.^33^ Due to that, increased levels of SAA1 and SAA2 proteoforms can be seen as a measure of the increased severity of the disease and so on prognostic factors. Due to its ubiquitous expression in several infectious diseases, SAA proteins cannot be associated directly with the SARS-CoV-2 and should be complemented with other viral specific molecular tests. A possible mechanism of increase of SAA proteins in severe COVID-19 patients could be due to the cytokine storm that is elicited during the infection. Indeed, increased levels of cytokines such as interleukin IL-2, IL-7, GCSF, interferon-γ inducible protein 10, MCP 1, MIP 1-α, and TNF-α and IL-6 is associated with COVID-19 disease severity, suggesting that the mortality observed could be due to virally/induced hyperinflammation.^34^ The elevation of IL-1 and IL-6 increase synergistically the levels of SAA proteins synergistically. At the same time, SAA proteins increase the expression of IL-1β mediated by NLRP3 in human and mouse immune cells.^35,36^ SARS-CoV ORF8b activates the NLRP3 inflammasome inducing the secretion of active IL-1β and IL-18.^37^ Moreover, SARS-CoV ORF3a activates the NLRP3 inflammasome by promoting TNF receptor-associated factor 3 (TRAF3)-ubiquitination of p105 and activation of NF-kB and subsequent transcription and secretion of IL-1β.^38^ Overactivation of NLRP3 in SARS-CoV-2 infection has been postulated delineating specific pathways for its activation^39–41^. Blockade of NFκB, a central player in the SAA-mediated activation of proinflammatory cytokines, could represent a novel therapeutic target for severe cases of COVID-19. Due to that, SAA proteins might play a critical role in SARS-CoV-2 infection as an early response to inflammation but also can be seen as pro-inflammatory proteins to amplify the cytokine storm. Although comprehensive LC-MS/MS analysis has been performed using sera from COVID-19 patients, a proteomic fingerprint using MALDI-TOF MS on plasma samples has not been reported. Recently, MALDI-TOF MS combined with a machine learning approach was used to detect SARS-CoV-2 in nasal swabs from infected patients.^42^ The application of RT-PCR, immunochromatography and recently MALDI-TOF MS have been used and proven to be reliable for the diagnosis of SARS-CoV-2 infection. However, no method exists so far to discriminate between high and low risk patients. This study shows that MALDI-TOF MS combined with machine learning algorithms offers a reproducible, easy to use, fast, low cost technique that can be implemented by several researchers worldwide to test the reliability of this marker. Moreover, the widespread use of MALDI-TOF in clinical laboratories will allow an easy transition into the hospitals.

### Limitations of the study

This study has focused on the fractionated plasma focusing on a limited mass range 2000-20000. Moreover, the concomitant ionization of proteins/peptides in this region limits the detection of low abundant ones. Improved large scale shotgun approaches combined with extensive fractionation have been applied to identify potential COVID-19 biomarkers and could be used in association with SAA1 and SAA2 provided in this study to create a panel of more reliable biomarkers. Association of the current biomarkers with other biomarkers will offer the possibility to improve the prognostic accuracy. Further validation in prospectively collected samples, as well as proof of added value to the existing noninvasive diagnostic strategies.

A larger independent cohort of patients should be analyzed to corroborate these findings. Inter-laboratory studies across countries should be performed to validate these data. Moreover, a time-course study during the development of the infection would give more information on the validity of these markers as early prognostic markers.

### Patient and public involvement

This study analyzed a retrospective case-series cohort. No patients were involved in the study design, setting the research questions, or the outcome measures directly. No patients were asked to give advice on interpretation or writing up of results.

## Data Availability

Raw data were submitted to PRIDE (https://www.ebi.ac.uk/pride/), project number PXD021581.

## Abbreviations

(ACN): Acetonitrile
(HCCA): Alpha-cyano-hydroxycinnamic acid
(AUC): Area under curve
(CV): Coefficient of variance
(COVID-19): Coronavirus disease 2019
(CRP): C-reactive protein
(DHB): Dihydroxybenzoic acid
(EDTA): Ethylenediamine tetra acetic acid
(IQR): Interquartile range
(MALDI-TOF MS): Matrix-assisted laser desorption ionization mass spectrometry
(PR): Precision-recall curve
(ROC): Receiver operating characteristic curve
(SAA): Serum amyloid A1/A2
(SA): Sinapinic acid
(SDS-PAGE): Sodium dodecyl sulfate–polyacrylamide gel electrophoresis
(TFA): Trifluoroacetic acid

## Acknowledgments

This work was supported by Fundação de Amparo à Pesquisa do Estado de São Paulo (FAPESP), GP (2018/18257-1, 2018/15549-1, 2020/04923-0), CW (2015/26722-8, 2017/03966-4), CRFM (2018/20468-0) and JCN (2020/04705-2). GP, CW, and CRFM were supported by Conselho Nacional de Desenvolvimento CientÍfico e Tecnológico (CNPq).

## Supplementary Figures

**Supplementary Figure 1:**
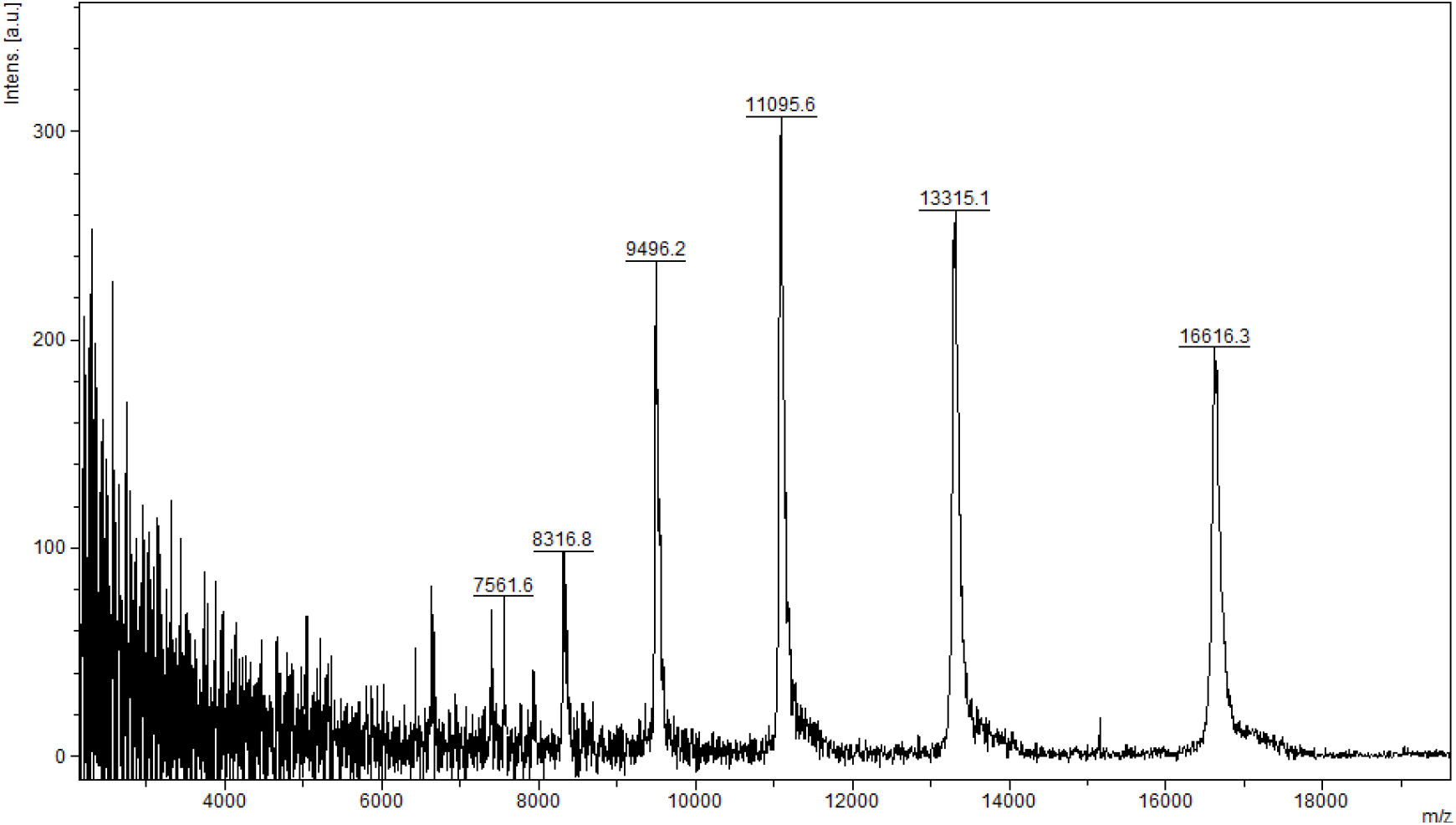
MALDI-TOF MS analysis in positive and linear mode of a pooled unfractionated plasma diluted (1:100) with HCCA (10mg/mL dissolved in acetonitrile 50%, water 49.9%, 0.1% TFA).

**Supplementary Figure 2:**
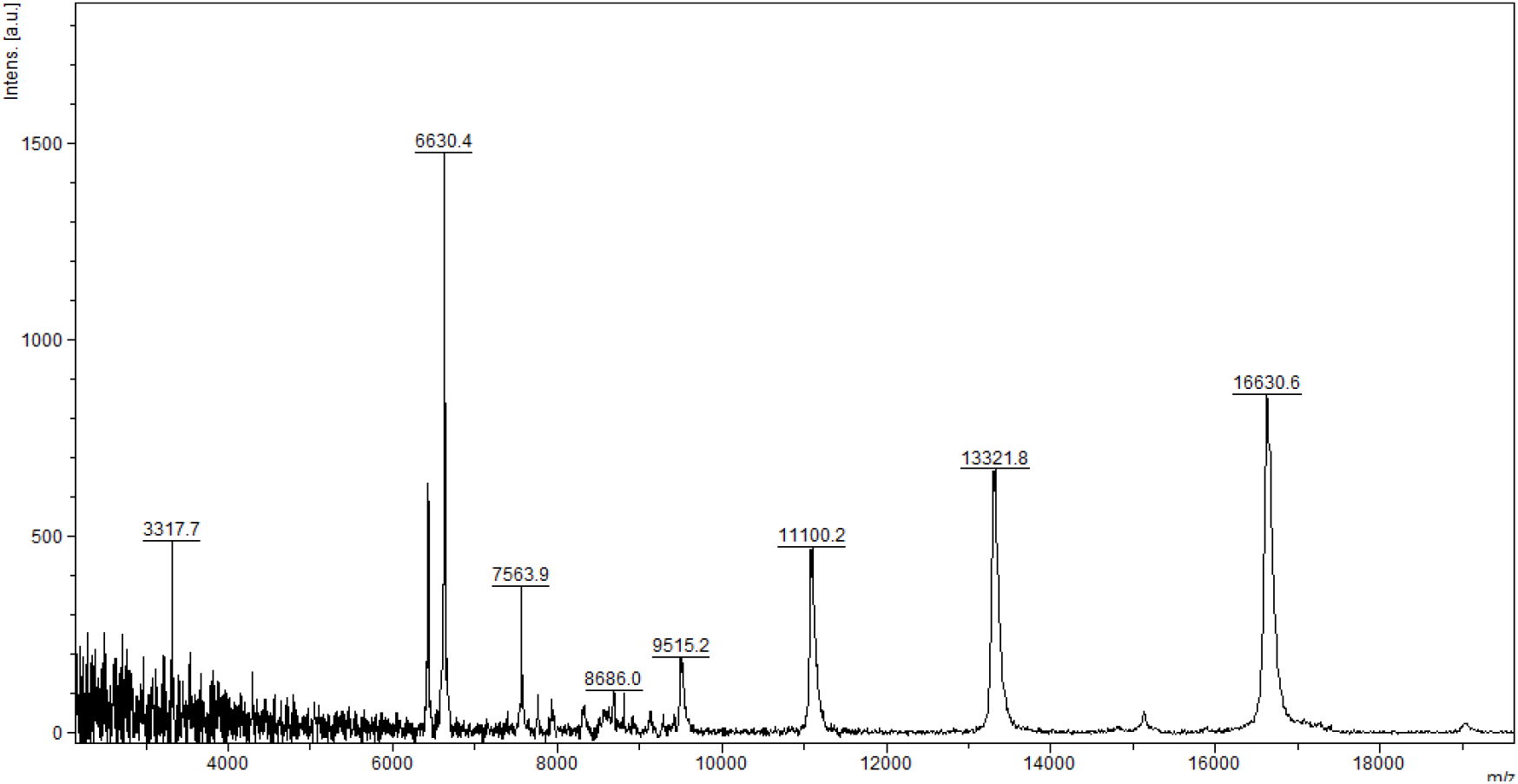
MALDI-TOF MS analysis in positive and linear mode of a pooled unfractionated plasma diluted (1:100) with HCCA (10mg/mL dissolved in acetonitrile 50%, water 47.5%, 2.5% TFA).

**Supplementary Figure 3:**
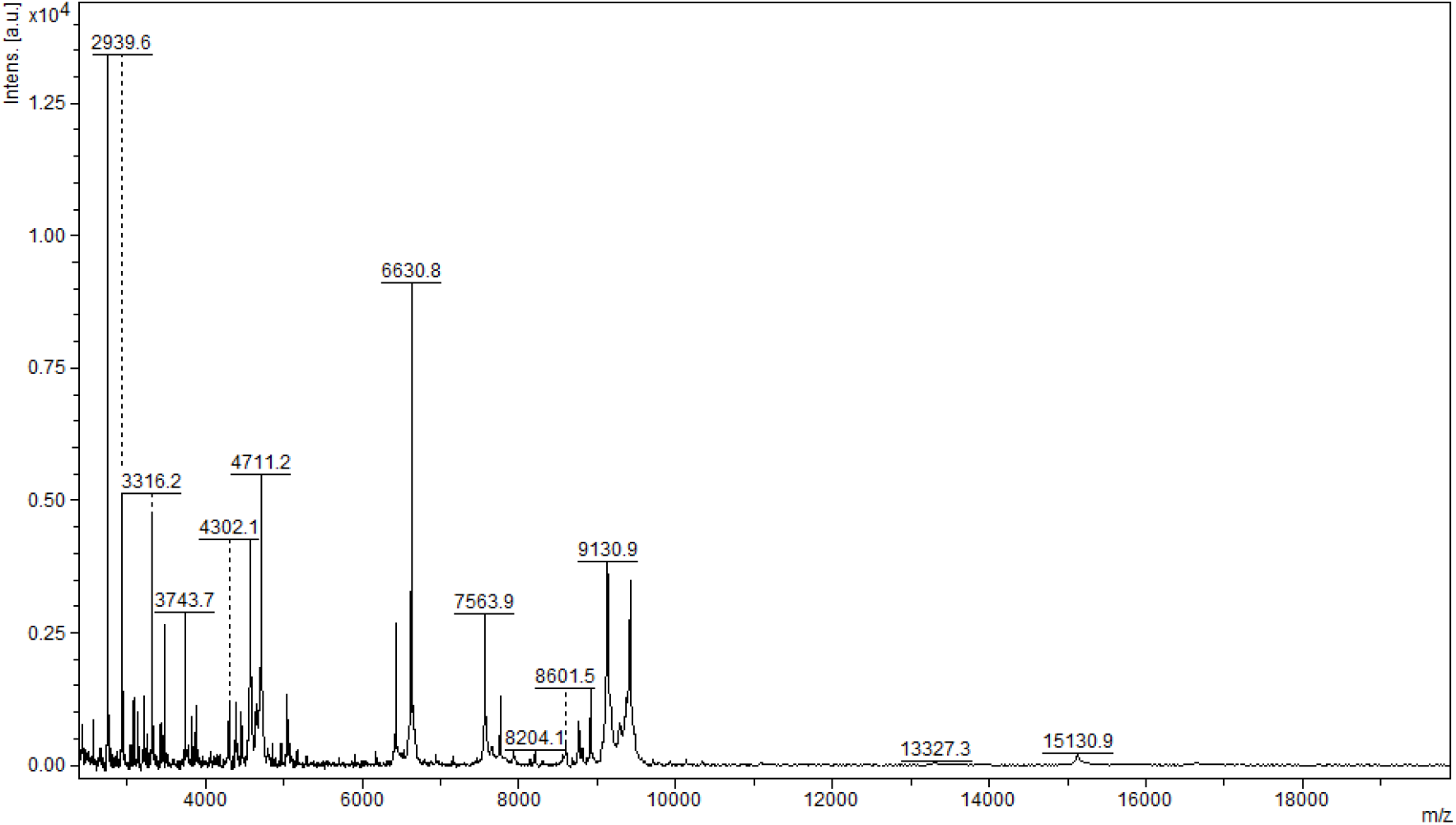
MALDI-TOF MS analysis in positive and linear mode of a microcolumn-based C18 fractionated plasma (1ul) analyzed with HCCA (10mg/mL dissolved in acetonitrile 50%, water 47.5%, 2.5% TFA).

**Supplementary Figure 4:**
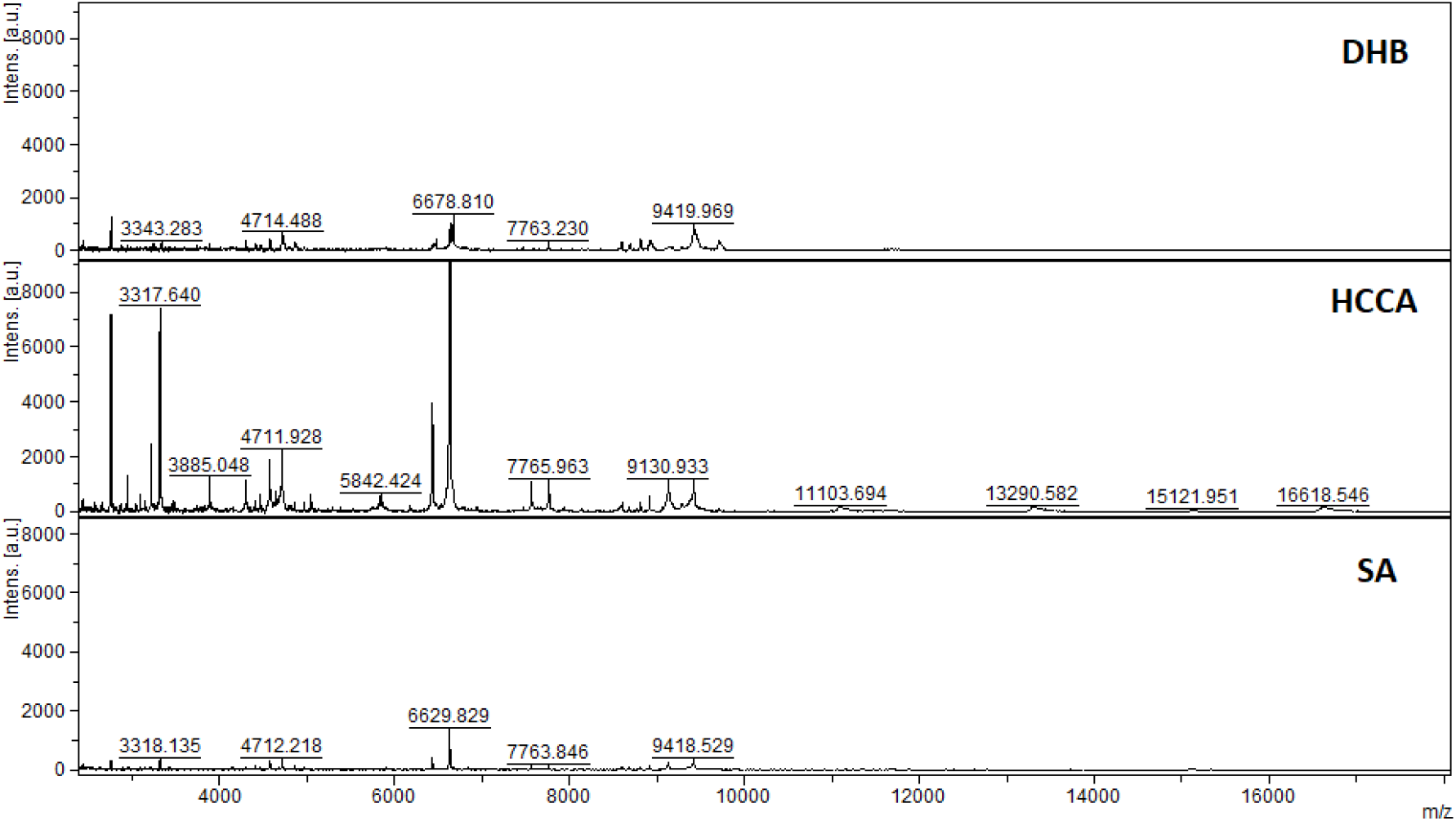
MALDI-TOF MS analysis in positive and linear mode of a microcolumn-based C18 fractionated plasma (1ul) analyzed with DHB, HCCA and SA (10mg/mL dissolved in acetonitrile 50%, water 47.5%, 2.5% TFA). Patient 199.

**Supplementary Figure 5:**
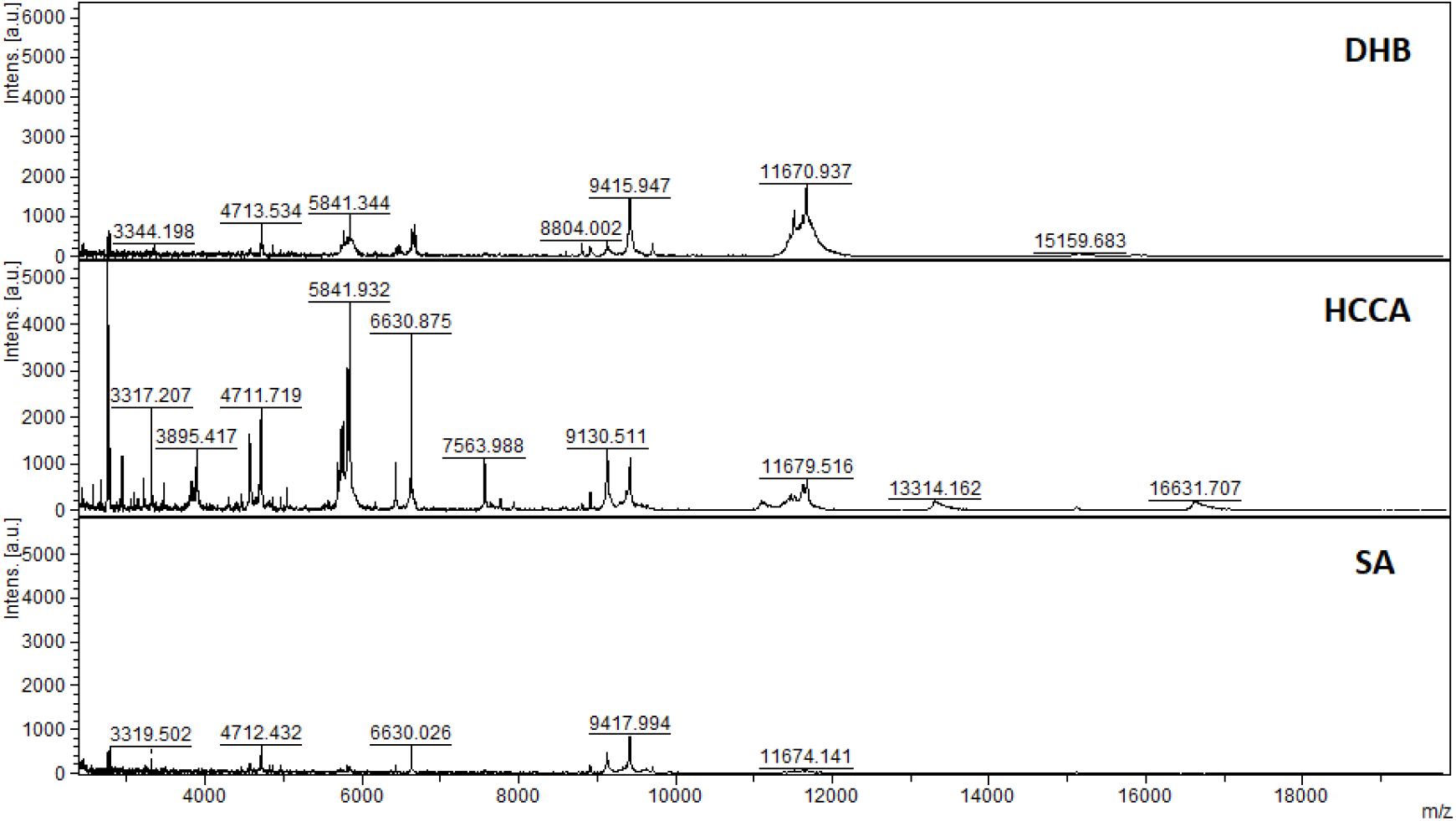
MALDI-TOF MS analysis in positive and linear mode of a microcolumn-based C18 fractionated plasma (1ul) analyzed with DHB, HCCA and SA (10mg/mL dissolved in acetonitrile 50%, water 47.5%, 2.5% TFA). Patient 198.

**Supplementary Figure 6:**
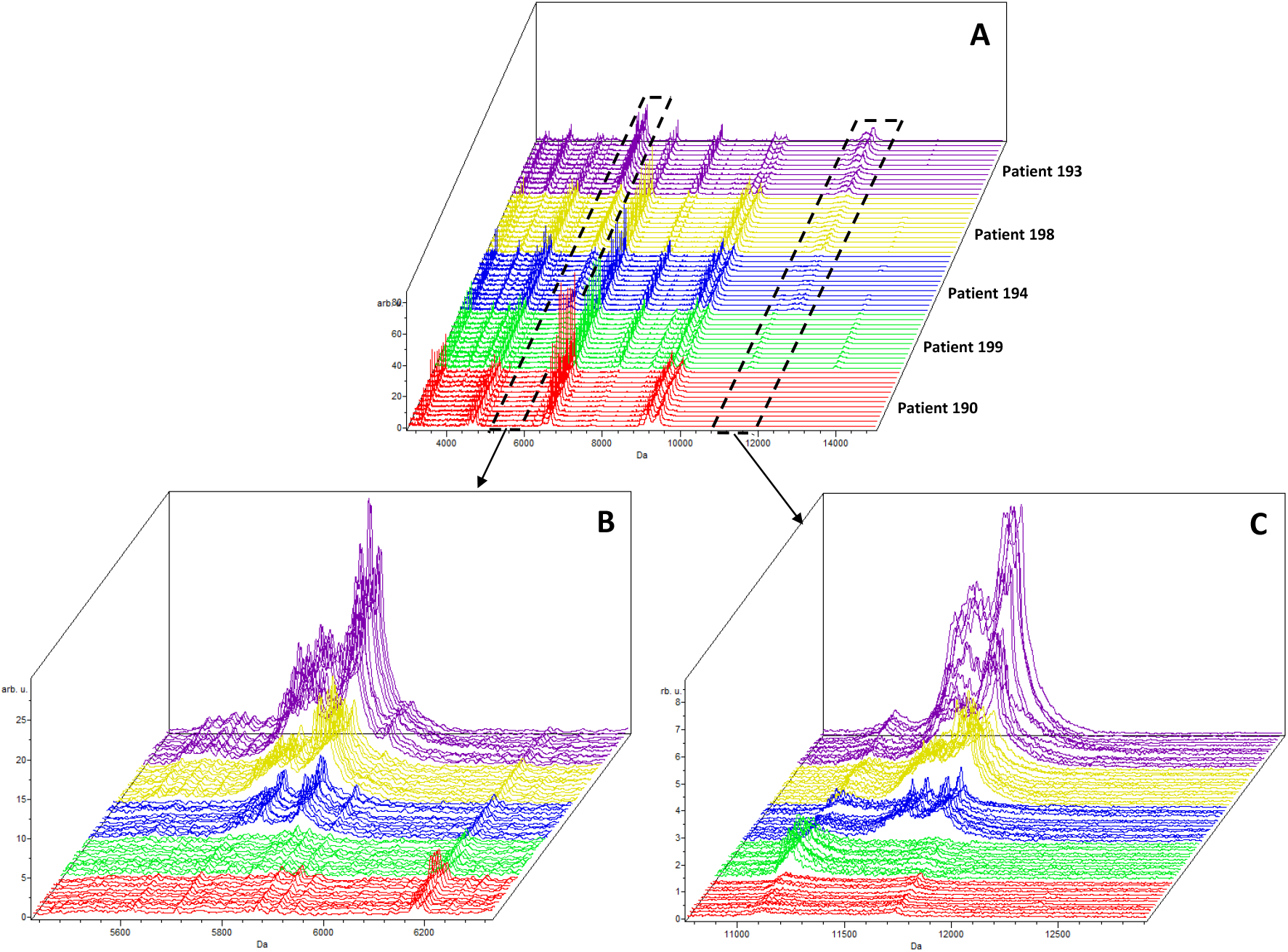
MALDI-TOF MS analysis in positive and linear mode of 5 patient plasma samples analyzed using 3 independent sample preparations and 4 acquisitions from each spot (total 12 spectra per patient). (A) Full range, (B) zoom at 5845 m/z and (C) zoom at 11683 m/z.

**Supplementary Figure 7:**
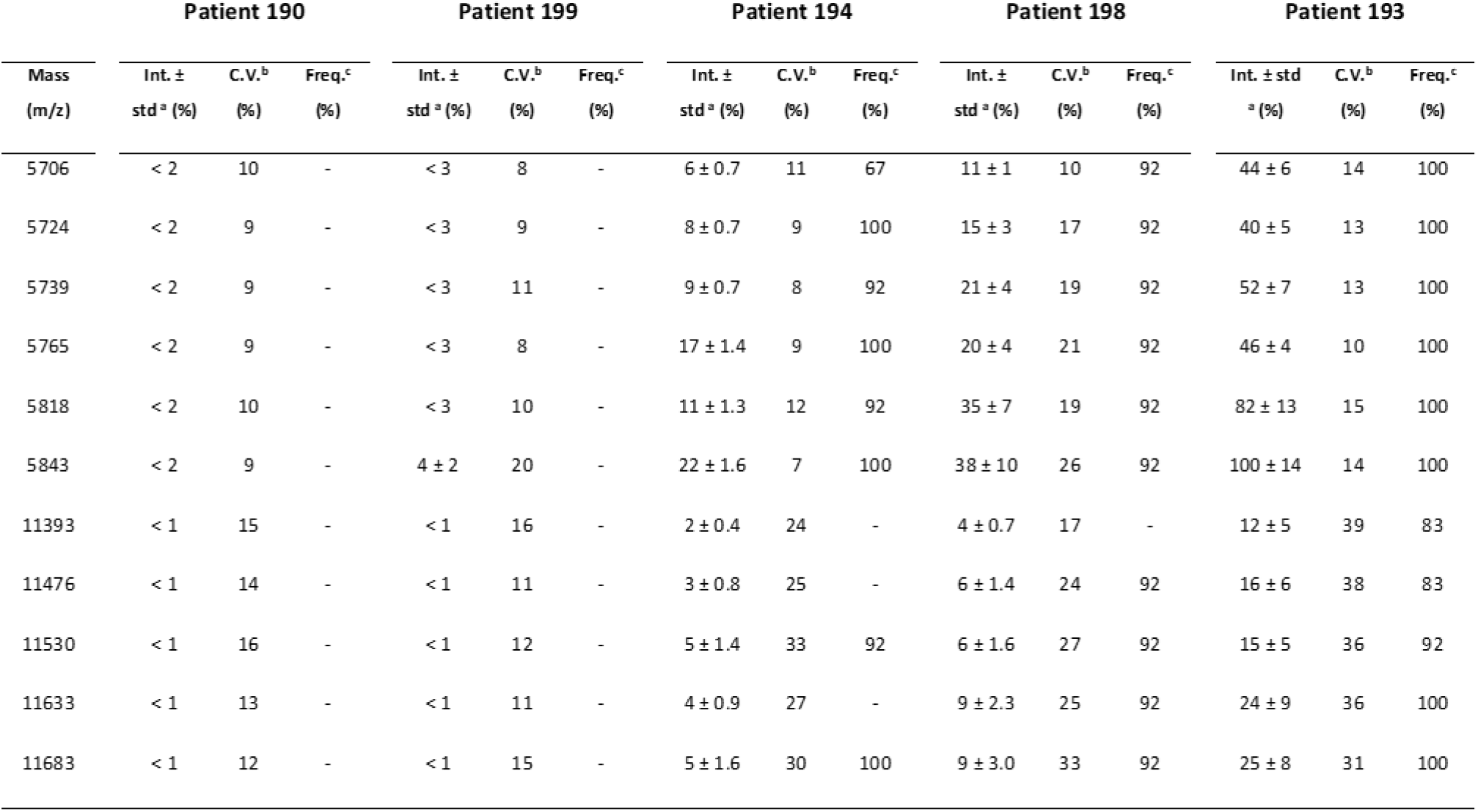
MALDI-TOF MS analysis in positive and linear mode of 5 patient plasma samples analyzed using 3 independent sample preparations and 4 acquisitions from each spot (total 12 spectra per patient). The intensities of peaks present in two selected mass regions, 5700-5900 and 11300-11700 m/z were analyzed. ^a^ Relative Intensity ± standard deviation; ^b^ Coefficient of Variation; ^c^ Peak Frequency - calculated from 12 spectra (3 prepared replicates and 4 spectra from each preparation) for peaks with a relative intensity > 5%.

**Supplementary Figure 8:**
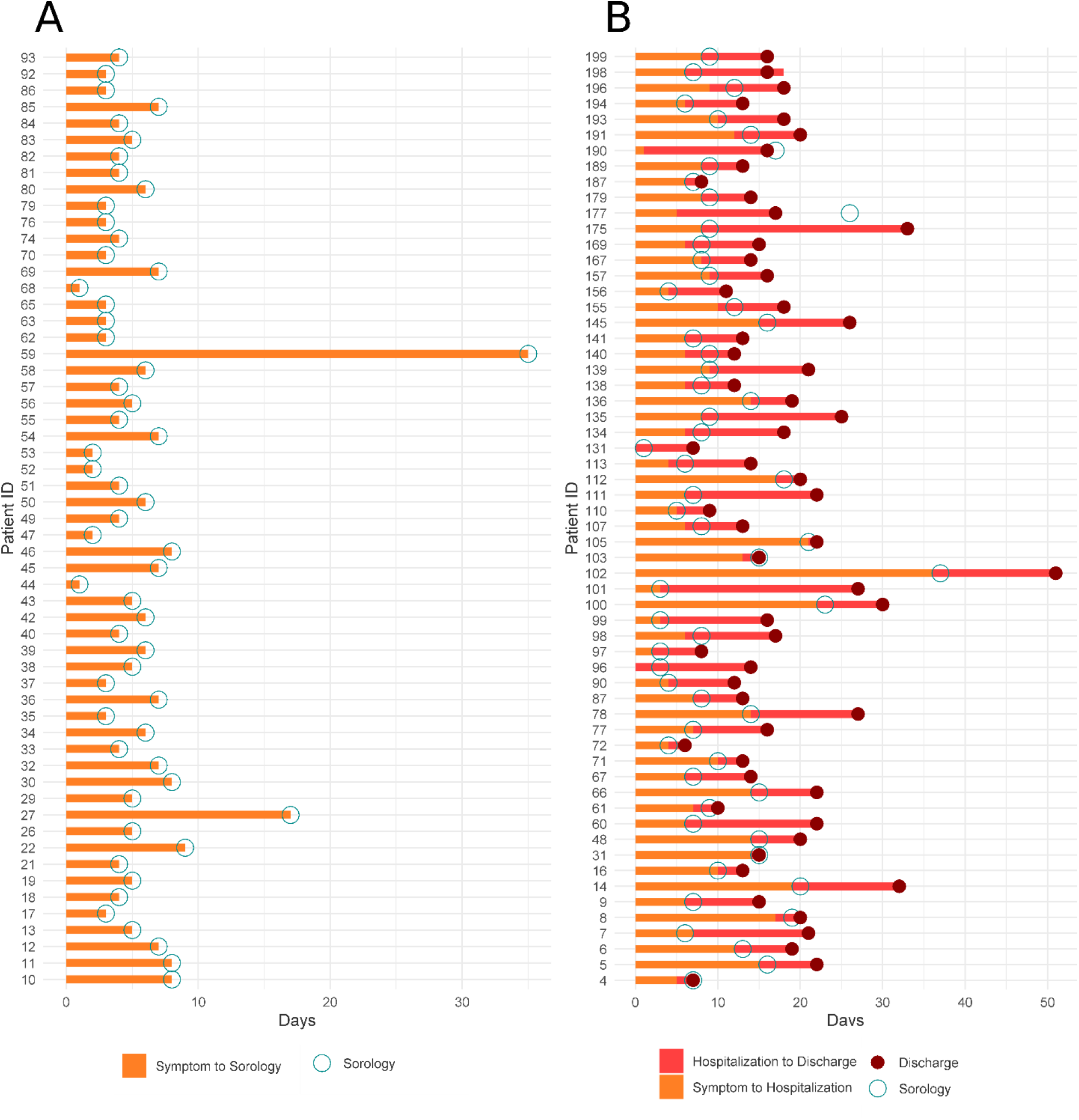
Timeline for patients from symptoms, admission to the hospital and discharge. The sample collection used for proteomics analysis is indicated.

## Supplementary Tables

Supplementary Table 1. Comorbidities associated with the 117 COVID-19 patients investigated in this study.

Supplementary Table 2: mass peaks (m/z) selected in the data preprocess step using Wilcoxon test and FSelector package.

Supplementary Table 3: Optimized parameters for each machine learning algorithm in the 4-fold Cross Validation.

Supplementary Table 4: Accuracy, Sensitivity and Specificity metrics of the 4-fold Cross Validation.

Supplementary Table 5: Average metrics for each machine learning algorithm tested with the non-filtered dataset.

Supplementary Table 6: Total proteins identified in the 10-15kDa region of C18 fractionated plasma collected from patients with high and low risk.

Supplementary Table 7: Total peptides identified in the 10-15kDa region of C18 fractionated plasma collected from patients with high and low risk.

